# Ivermectin as a potential treatment for mild to moderate COVID-19 – A double blind randomized placebo-controlled trial

**DOI:** 10.1101/2021.01.05.21249310

**Authors:** Ravikirti, Ranjini Roy, Chandrima Pattadar, Rishav Raj, Neeraj Agarwal, Bijit Biswas, Pramod Kumar Majhi, Deependra Kumar Rai, Shyama, Anjani Kumar, Asim Sarfaraz

**Author notes:** **Corresponding Author** Dr Ravikirti, Additional Professor and Head, Department of General Medicine, All India Institute of Medical Sciences (AIIMS), Patna, India.

## Abstract

**Objective:** Ivermectin has been suggested as a treatment for COVID-19.This randomised control trial was conducted to test the efficacy of Ivermectin in the treatment of mild and moderate COVID-19.

**Design:** Parallel, double blind, randomised, placebo controlled trial Setting: A tertiary care dedicated COVID-19 hospital in Bihar, India

**Participants:** Adult patients (> 18 years) admitted with mild to moderate COVID 19 disease (saturation > 90% on room air, respiratory rate < 30 and no features of shock) with no contraindications to ivermectin and willing to participate in the study

**Intervention:** Patients in the intervention arm were given ivermectin 12 mg on day 1 and day 2 of admission. Patients in the placebo arm were given identical looking placebo tablets. Rest of the treatment was continued as per the existing protocol and the clinical judgment of the treating teams.

**Outcome Measures:** The primary outcome measure was a negative RT-PCR test for SARS-CoV-2 on day 6 of admission. The secondary outcome measures were symptom status on day 6, discharge status on day 10, admission to ICU, need for invasive mechanical ventilation and in-hospital mortality.

**Results:** A total of 115 patients were enrolled for the study of which 112 were included in the final analysis. Of them, 55 were randomised to the intervention arm while 57 were randomised to the placebo arm. There was no significant difference in the baseline characteristics of the two arms. There was no significant difference in the primary outcome, i.e. negative RT-PCR status on day 6 between the two groups. Similarly, there was no significant difference between the two groups in most of the secondary outcome measures, viz. symptom status on day 6, discharge status on day 10, admission to ICU, and need for invasive mechanical ventilation. However, while there was no in-hospital mortality in the intervention arm, there were 4 deaths in the placebo arm. As a result, all patients in the intervention arm (n=56) were successfully discharged as compared to 93.1% (n=54/58) in the placebo arm (RR 1.1, 95% CI 1.0 to 1.2, p=0.019).

**Conclusion:** There was no difference in the primary outcome i.e. negative RT-PCR status on day 6 of admission with the use of ivermectin. However, a significantly higher proportion of patients were discharged alive from the hospital when they received ivermectin.

**Strengths and Limitations of the Study:**

- This study was randomised and double blind, thereby minimizing the chance of bias.
- All outcome measures except symptom status on day 6 were objective and placebo control was used for comparison.
- Only single repeat RT-PCR was done. So median time to viral clearance in the two groups could not be calculated.
- Severe cases were not included in the study.

## Introduction

The SARS-CoV-2 infection first reported from Wuhan in China in November 2019 has grown into a pandemic affecting almost all regions of the world. More than eighty six million people have been affected worldwide with over 1.8 million deaths reported already.^1^ The case fatality rate has varied from less than one percent in some countries to 18.3% in the Lombardy region of Italy.^2^ Though several treatments have been tried, none has been convincingly found to be effective except low dose steroids in patients requiring supplemental oxygen or mechanical ventilation.^3, 4^ Hydroxychloroquine was used widely after a small French study at the beginning of the pandemic found that 20 patients who received the drug with or without Azithromycin showed greater viral clearance on day 6 compared to those who did not receive it.^5^ However, no significant benefit was noted in larger and better designed studies.^6, 7^ Lopinavir/Ritonavir, a combination used in the treatment of HIV and repurposed for use in COVID-19 failed to show any significant benefit in hospitalized adult patients in a randomized controlled open-label trial of 199 patients.^8^ Like hydroxychloroquine, the SOLIDARITY trial did not find any meaningful benefit from any of the other repurposed antiviral drugs, viz. lopinavir, interferon (given with lopinavir) and remdesivir.^7^ Remdesivir, however, has been shown to shorten the median recovery time in another randomized double-blind, placebo-controlled trial.^9^ Tocilizumab, an interleukin-6 receptor blocker, expected to be effective in dampening the cytokine storm that some patients experience was not found to be effective in preventing intubation or death in moderately ill hospitalized patients with Covid-19^10^. Similarly, studies have not demonstrated any benefit from using convalescent plasma of patients who have recovered from the disease. E.g. an open label, parallel arm, phase-II, multicentric, randomised controlled trial of 464 adults with moderate COVID-19 concluded that convalescent plasma was not associated with a reduction in progression to severe disease or all-cause mortality.^11^

There has been a growing interest in the anti-parasitic drug, Ivermectin ever since it was reported to have an in-vitro activity against SARS-CoV-2.^12^ The drug belongs to a class of macrocyclic lactones called the avermectins that induce paralysis by activating a family of ligand-gated Cl^−^ channels, particularly glutamate-gated Cl^−^ channels found in invertebrates.^13^ It is the drug of choice for onchocerciasis in adults and children above 5 years of age. Additionally, it is used to treat lymphatic filariasis, strongyloidiasis, cutaneous larva migrans and scabies.^14^ For most helminthic infestations it is administered as a single oral dose of 150–200 μg/kg. The dose may be repeated after 24 hours in intestinal strongyloidiasis. Peak level in plasma is achieved in four to five hours after oral administration. It has a long half-life of 57 h in adults, is about 93% bound to plasma proteins, is extensively metabolized by hepatic CYP3A4 and is not excreted in the urine.^15^ Its anti-viral activity, particularly against RNA viruses, is believed to be through the inhibition of nuclear import of several host and viral proteins.^12^

Several observational studies have reported a beneficial effect of Ivermectin in COVID-19.^16, 17, 18, 19, 20^ However, other studies including a large retrospective review of 5683 patients from Peru receiving different drug combinations have found no benefit.^21, 22^

Many interventional studies have also reported positive outcomes lately. In a non-randomised study where 16 patients with mild to moderate disease were given a single dose of Ivermectin 200 mcg/kg in addition to the standard treatment with hydroxychloroquine had a shorter time to recovery compared to a synthetic controlled arm of 71 patients who did not receive the drug.^23^ A small open label proof of concept study with voluntary allocation noted that all the 28 patients with mild to moderate disease who received the combination of ivermectin (6 mg once daily on days 0, 1, 7 and 8), azithromycin (500 mg once daily for 4 days) and cholecalciferol (4000 UI twice daily for 30 days) in addition to standard treatment were RT-PCR negative on day 10 while all the 7 patients in the control group remained RT-PCR positive.^24^ Another open label trial of 140 COVID-19 patients with group allocation based on the date of recruitment noted a shorter time to recovery and improved survival in severe disease with the use of ivermectin 200 mcg/kg daily for two days (A third dose was given to some patients on day 7.) along with doxycycline 100 mg twice daily for 5 to 10 days compared to patients who received standard care alone. ^25^ A trial of 72 patients that compared oral ivermectin 12 mg daily for 5 days, a single dose of ivermectin 12 mg with doxycycline 200 mg stat followed by 100 mg twice daily for four days and placebo noted a quicker viral clearance with a five day course of ivermectin but not with the regime that combined it with doxycycline.^26^ A yet to be published RCT of 400 patients has found a lower rate of progression to severe disease and higher rates of clinical recovery and viral clearance with the use of a single dose of ivermectin 12 mg and doxycycline 100 mg twice daily for 5 days.^27^

On the other hand, an open label RCT of 116 failed to show any statistically significant difference in the time to symptomatic recovery or virological clearance as evidenced by a negative RT-PCR test between the group that received a single dose of Ivermectin 200 mcg/kg along with doxycycline 100 mg twice daily for 10 days and the one that received hydroxychloroquine 400 mg on day 1 followed by 200 mg twice daily for 10 days along with azithromycin 500 mg daily for 5 days.^28^ Similarly, a prospective comparative study of 400 patients with mild to moderate disease comparing ivermectin 18 mg on day 1 and doxycycline 100 mg twice daily for 5 days (Group A) with hydroxychloroquine 800 mg on day 1 followed by 400 mg daily for 10 days and Azithromycin 500 mg on day 1 followed by 250 mg daily for 4 days (Group B) did not find any improvement in viral clearance with the Ivermectin and doxycycline combination.^29^ Several smaller RCTs have reported no benefit with ivermectin.^30, 31, 32, 33^ A trial of 585 patients concluded that combined with azithromycin, the use of nitazoxanide, ivermectin, and hydroxychloroquine led to similar clinical outcomes which were better than those in a retrospective control group of patients who had not received any of these three drugs.^34^

Here, we report the results of a double blind RCT comparing Ivermectin 12 mg on two consecutive days with placebo in patients with mild to moderate COVID-19.

## Materials and Methods

The aim of the study was to assess the efficacy of ivermectin in the management of mild to moderate COVID 19. The study design was double blind randomized placebo-controlled trial. Ethical clearance was obtained from the Institutional Ethics Committee, All India Institute of Medical Sciences (AIIMS), Patna. The trial was registered with the Clinical Trials Registry – India (registration number – CTRI/2020/08/027225). Patients or the public could not be involved in the design, conduct, reporting or dissemination planning of this study.

Assuming an improvement of 30% in 10 day recovery in patients receiving the intervention (50%) compared to the standard of care (20%) with 5% absolute precision and 80% power the total sample size was calculated to be 90 using OpenEpi software with equal allotment in both the arms.

All patients above the age of 18 admitted with a diagnosis of COVID -19 (on the basis of a positive RT-PCR or Rapid Antigen Test report) at AIIMS, Patna, India with mild or moderate disease as defined by the ministry of health and family welfare guidelines (Table 1) and not meeting any of the exclusion criteria were considered eligible for the study. The exclusion criteria were: known allergy to or adverse drug reaction with Ivermectin; unwillingness or inability to provide consent to participate in the study; prior use of ivermectin during the course of this illness; pregnancy and lactation.

**Table 1:**
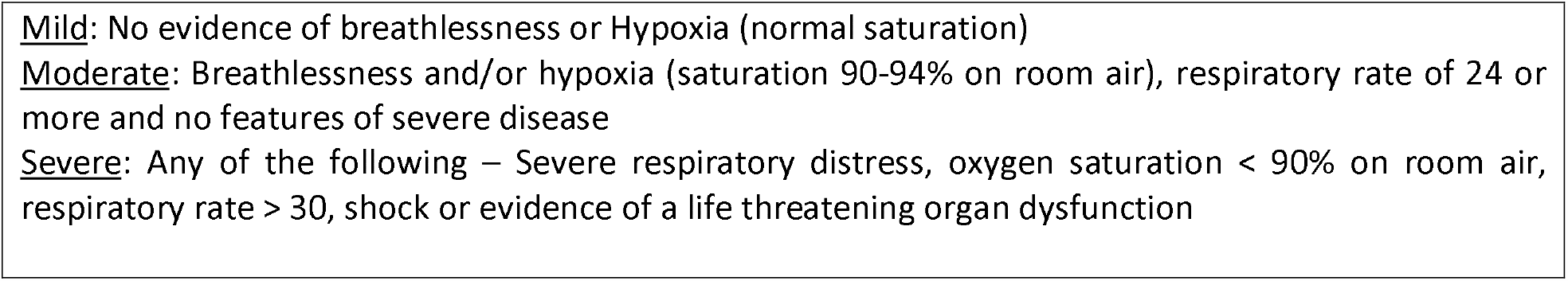
Definitions of Mild, moderate and severe COVID-19^35^.

|  |
| --- |
| <u>Mild</u>: No evidence of breathlessness or Hypoxia (normal saturation) |
| <u>Moderate</u>: Breathlessness and/or hypoxia (saturation 90-94% on room air), respiratory rate of 24 or more and no features of severe disease |
| <u>Severe</u>: Any of the following – Severe respiratory distress, oxygen saturation < 90% on room air, respiratory rate > 30, shock or evidence of a life threatening organ dysfunction |

All patients admitted between 1 August and 31 October 2020, meeting the eligibility criteria and willing to participate were included in the study. After obtaining informed consent, they were randomly assigned to either group A or group B. Block randomisation was done with variable block sizes of 4, 6 and 8. A unique random allocation table was generated using the Sealed Envelope software. Once a patient had consented to participate in the study, they were allocated an envelope as per the sequence, assigning them to one of the two groups. The person doing the randomisation was not a part of the investigating team. One of these two groups was the intervention group and the other was the placebo group. However, up until the analysis of the data, this information was confined to the pharmacist dispensing the tablets.

The patients’ baseline characteristics like age, sex, comorbidities, duration of symptoms and disease severity on admission were noted down at the time of recruitment. Their clinical status including heart rate, blood pressure, respiratory rate and oxygen saturation were recorded every day during their stay in the hospital. All patients were managed by their treating teams according to the standard guidelines and their clinical judgment.^36^ In addition, patients in the intervention arm were given ivermectin 12 mg on days 1 and 2. The patients in the placebo arm received identical looking placebo tablets. All patients had to have a second RT-PCR done on Day 6. Any adverse reactions and symptom status (asymptomatic or still symptomatic) on Day 6 were also noted.

Discharge decisions were made by the respective treating teams, according to the existing protocol. The discharge criteria were: 1. Ten days from the onset of symptoms, 2. Afebrile for three days and 3. Maintaining O2 saturation above 94% without supplemental oxygen for four days.

The following outcomes were measured:

Primary outcome: A negative RT-PCR report on day 6.

Secondary outcomes:

1. Whether or not symptomatic on day 6
2. Discharge by day 10
3. Admission to ICU
4. Need for invasive mechanical ventilation
5. In-hospital mortality

Statistical analysis was done using IBM SPSS (Chicago, USA) software, version-22. All descriptive data were expressed as mean (SD) and frequency (percentage). Bivariate analysis was performed using the independent samples ‘t’ test for continuous variables and Fishers Exact test for categorical variables. Strengths of association between the two groups and various outcome variables, viz. RT-PCR result on day 6, symptom status on day 6, discharge status on day 10, ICU requirement, need for mechanical ventilation and in-hospital mortality were reported as rate ratios (RR). The minimum acceptable confidence level was α=0.95 for all data and any difference with p<0.05 was considered significant.

## Results

A total of 115 patients were enrolled for the study (Figure 1). 57 were randomised to the intervention arm while 58 were randomised to the placebo arm. One patient in either arm was administered ivermectin by the treating team and one patient in the intervention arm was lost to follow up from day 2. Excluding these three patients, 55 patients in the intervention arm and 57 patients in the placebo arm were included in the final analysis. Table 2 shows baseline characteristics of the two arms including age, sex, comorbidities, disease severity and the treatments given while in the hospital. There was no significant difference in any of the variables. Table 3 shows the primary and secondary outcomes in the two groups. 23.6% of the patients (n=13) in the intervention arm and 31.6 % (n=18) in the placebo arm tested RT-PCR negative for SARS-CoV-2 on day 6. The difference was not significant (RR 0.8, 95% CI 0.4 to 1.4, p=0.347). 83.6% (n=46) of the patients in the intervention arm and 89.5% (n=51) in the placebo arm were asymptomatic by day 6 (RR 0.9, 95% CI 0.8 to 1.1, p=0.363). 80% (n=44) and 73.7% (n=42) of the patients in the two arms respectively had been discharged by day 10 (RR 1.2, 95% CI 0.7 to 1.9, p=0.428). 9.1% (n=5) of the patients in the intervention arm and 10.5% (n=6) in the placebo arm required ICU care (RR 0.9, 95% CI 0.3 to 2.7, p=0.798). Only 1.8% (n=1) in the intervention arm needed invasive ventilation compared to 8.8% (n=5) in the placebo arm. However, the difference did not reach statistical significance (RR 0.2, 95% CI 0 to 1.7, p=0.088). In-hospital mortality was 6.9% (n=4) in the placebo arm as opposed to no deaths in the intervention arm. Conversely, all patients in the intervention arm (n=55) were successfully discharged as compared to 93% (n=53) in the placebo arm (RR 1.1, 95% CI 1.0 to 1.2, p=0.019).

**Table 2:** Background Characteristics of the study subjects:

| Variable | Ivermectin<br>Group<br>55 (49.1%)<br>N (%) /<br>Mean±SD | Placebo<br>Group<br>57(50.9%)<br>N (%) /<br>Mean±SD | Total<br>112 (100%)<br>N (%) /<br>Mean±SD | Chi-Square /<br>'t' Value | p-<br>value |
| --- | --- | --- | --- | --- | --- |
| **Age in years:** | 50.7±12.7 | 54.2±16.3 | 52.5±14.7 | 1.239 | .218* |
| **Gender:** |  |  |  |  |  |
| Female | 15 (27.3) | 16 (28.1) | 31 (27.7) | .009 | 1.00<sup>#</sup> |
| Male | 40 (72.7) | 41 (71.9) | 81 (72.3) |  |  |
| **Days since onset of symptoms:** | 6.1±3.6 | 7.9±8.6 | 6.9±6.6 | 1.443 | .152* |
| **Comorbidities:** |  |  |  |  |  |
| Hypertension | 21 (38.2) | 18 (31.6) | 39 (34.8) | .538 | .553<sup>#</sup> |
| Diabetes | 21 (38.2) | 19(33.3) | 40 (35.7) | .287 | .694<sup>#</sup> |
| Ischemic Heart Disease | 2 (3.6) | 8 (14.0) | 10 (8.9) | 3.722 | .094<sup>#</sup> |
| Heart Failure | 1 (1.8) | 1 (1.8) | 2 (1.8) | .001 | 1.00<sup>#</sup> |
| Chronic Kidney Disease | 1 (1.8) | 2 (3.5) | 3 (2.7) | .307 | 1.00<sup>#</sup> |
| Stroke | 0 (0.0) | 0 (0.0) | 0 (0.0) | - | - |
| Chronic Obstructive Pulmonary<br>Disease | 1 (1.8) | 0 (0.0) | 1 (0.9) | 1.046 | .491<sup>#</sup> |
| Asthma | 1 (1.8) | 0 (0.0) | 1 (0.9) | 1.046 | .491<sup>#</sup> |
| Cancer | 2 (3.6) | 4 (7.0) | 6 (5.4) | .631 | .679<sup>#</sup> |
| Other comorbidities | 7 (12.7) | 11 (19.3) | 18 (16.1) | .896 | .443<sup>#</sup> |
| **COVID-19 disease severity on**<br>**admission:** |  |  |  |  |  |
| Mild | 42 (76.4) | 46 (80.7) | 88 (78.6) | .313 | .648<sup>#</sup> |
| Moderate | 13 (23.6) | 11 (19.3) | 24 (21.4) |  |  |
| **Treatments given:** |  |  |  |  |  |
| Hydroxychloroquine | 55 (100.0) | 57 (100.0) | 112 (100.0) | - | - |
| Steroid | 55 (100.0) | 57 (100.0) | 112 (100.0) | - | - |
| Enoxaparin | 53 (96.4) | 55 (96.5) | 108 (96.4) | .001 | 1.00<sup>#</sup> |
| Antibiotics | 55 (100.0) | 57 (100.0) | 112 (100.0) | - | - |
| Remdesivir | 12 (21.8) | 11 (19.3) | 23 (20.5) | .109 | .817<sup>#</sup> |
| Convalescent Plasma | 8 (14.5) | 7 (12.3) | 15 (13.4) | .124 | .786<sup>#</sup> |
| Tocilizumab | 4 (7.3) | 3 (5.3) | 7 (6.3) | .193 | .714<sup>#</sup> |
| Other Drugs | 36 (65.5) | 38 (66.7) | 74 (66.1) | .018 | 1.00<sup>#</sup> |

\*Independent Samples 't' Test; <sup>#</sup>Fishers Exact Test

**Table 3:** Distribution of the study subjects according to the primary and secondary outcomes.

| Outcome Variable | Ivermectin Group<br>(N=55)<br>N (%) | Placebo Group<br>(N=57)<br>N (%) | Rate Ratio (RR)<br>(95% CI) | P value* |
| --- | --- | --- | --- | --- |
| **Primary Outcome** |  |  |  |  |
| Negative RT-PCR on day 6 | 13 (23.6) | 18 (31.6) | 0.8 (0.4-1.4) | .347 |
| **Secondary Outcomes** |  |  |  |  |
| Symptom free on day 6 | 46 (83.6) | 51 (89.5) | 0.9 (0.8-1.1) | .363 |
| Discharged by day 10 | 44 (80.0) | 42 (73.7) | 1.2 (0.7-1.9) | .428 |
| Admission to ICU | 5 (9.1) | 6 (10.5) | 0.9 (0.3-2.7) | .798 |
| Invasive Ventilation | 1 (1.8) | 5 (8.8) | 0.2 (0.0-1.7) | .088 |
| Final outcome: discharge | 55 (100.0) | 53 (93.0) | 1.1 (1.0-1.2) | .019 |
| Final outcome: in-hospital mortality | 0 (0) | 4 (7) | 0 | - |

RR: Rate Ratio, CI: confidence interval; \* Fishers Exact Test

**Figure 1:**
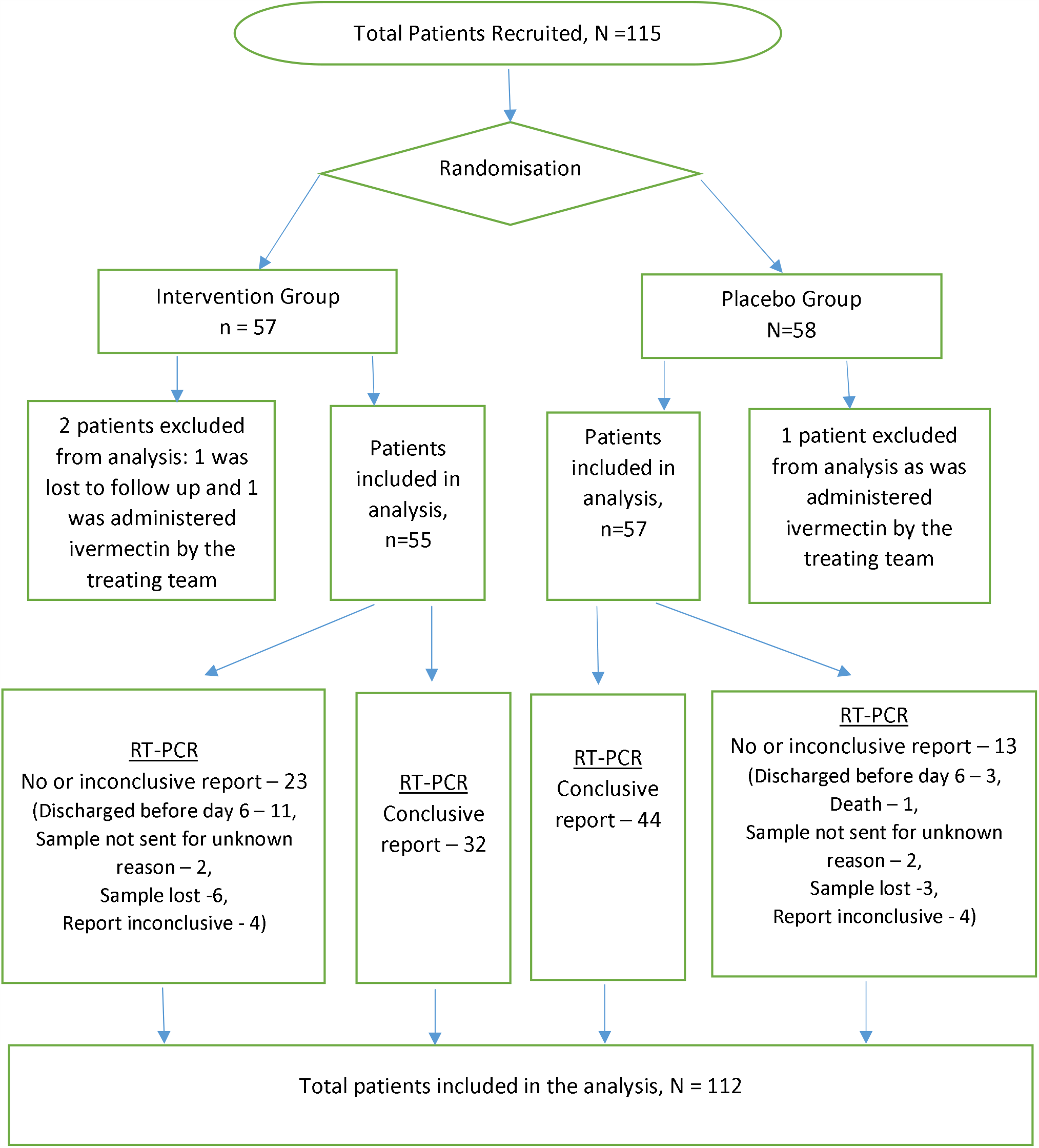
Flow-chart of the clinical trial.

## Discussion

This study did not find any benefit with the use of ivermectin in the primary outcome i.e. negative RT-PCR on day 6 as well as in most of the secondary outcomes, viz. symptom status on day 6, discharge by day 10, admission to ICU and use of invasive ventilation in mild and moderate COVID-19. However, there was no in-hospital mortality in the intervention arm leading to a significantly higher chance of being discharged alive from the hospital.

Some of the previous observational studies^16, 19^, a non-randomised interventional study^24^ and some RCTs^26, 27^ have reported quicker viral clearance with the use of ivermectin. However, other studies have not found this benefit^22, 28^.There was variation in the regimes used by these studies. Notably, in the current study a conclusive RT-PCR report could not be obtained in 36 (32.1%) out of the 112 patients (figure 1). The sample could not be sent in 19 patients as 1 patient died and 14 patients got discharged before day 6 while the reason was not clear in 4 patients. The sample got lost in 9 patients while there was inconclusive report in 8 patients. Furthermore, a single repeat RT-PCR was done on day 6 as opposed to serial testing.

It has been suggested that prolonged viral RNA shedding may continue for weeks after recovery leading to a false positive RT-PCR report even though the patient may not be infective any more.^37^ Thus, it is questionable whether a negative repeat RT-PCR is a clinically useful outcome measure.

The only significant positive outcome of the current study was improved survival with the use of ivermectin. This was also noted earlier in some of the observational studies^18, 19, 20^. Similarly, an open label RCT of 140 patients found lower mortality with a combination of ivermectin and doxycycline although the difference did not reach statistical significance^25^. A larger, yet to be published, RCT has reported a mortality of 1.67% in the placebo group compared to 0% in the group that was given ivermectin and doxycycline^27^.

It may appear surprising that all patients in the current trial received corticosteroids even though 78.8 % of the patients had only mild disease (table 2). This is because the first dose was prescribed by the doctor on duty in all patients. However, the drug was stopped on the subsequent consultant round in most patients with mild disease.

The findings of this trial should be interpreted with consideration to certain limitations. As mentioned above, a conclusive repeat RT-PCR report could not be obtained in 32.1% of the patients. Moreover, as serial RT-PCR tests were not done, the median time to viral clearance in the two groups could not be ascertained. As only mild and moderate cases were included, it cannot be said whether the benefit in survival seen with ivermectin can also be seen in severe cases. Similarly, it cannot be said on the basis of this study whether or not using higher doses of ivermectin or combining it with other drugs like doxycycline would offer any additional benefit. Similar but larger studies may be able to give a more definitive answer, especially in relation to the other secondary outcome measures.

## Data Availability

The data can be made available by the corresponding author on rasonable request.

## Acknowledgements

We would like to thank Dr PK Singh, Director AIIMS Patna for facilitating the study, Dr Alok Ranjan, Assistant Professor, department of Community and Family Medicine, AIIMS, Patna for reviewing the statistical analysis, Mr Sunil Verma of Sun Pharma Pvt. Ltd. for arranging the placebo tablets and Mr Ambuj Kumar, Pharmacist at AIIMS, Patna for dispensing the tablets and ensuring blinding.

